# Establishing Reference Values for Peripheral Blood Neutrophil CD64 index and Monocyte HLA-DR of Healthy Adult

**DOI:** 10.1101/2025.10.29.25339107

**Authors:** Liangjun Zhang, Yi Li, Huixiu Zhong, Jingyuan Huang, Minggang Yin

## Abstract

Neutrophil CD64 (nCD64) index and Monocyte HLA-DR (mHLADR) played an important role in the diagnosis of infection, but their levels in healthy adults have not been thoroughly investigated. This study established reference values for nCD64 index and mHLADR% in the peripheral blood from 285 healthy adults. All the subjects were grouped into subgroups according to sex (male and female) and age (14–51 years, 51–100 years). The analyses indicated age to be an important factor associated with changes in nCD64 index, which gradually increased from 14 to 100 years and showed a positive correlation with age (P < 0.0001). In contrast, no significant differences in mHLA-DR% were observed across gender or age groups (p > 0.05). Furthermore, no correlation was found between nCD64 index or mHLA-DR% and all the inflammatory derived indicators of blood routine (P > 0.05). In conclusion, the reference intervals established for the nCD64 index and mHLA-DR% in healthy adults may provide supportive information for monitoring immune status in the context of infectious diseases..

## 1. Introduction

Neutrophil CD64(nCD64) is a cell surface transmembrane glycoprotein with high affinity for IgG and is a member of the immunoglobulin family, also known as Fc segment type 1 receptor (FC-γR1), which is mainly expressed on macrophages, monocytes and dendritic cell^1^, while it is weakly expressed on the surface of neutrophils. Upon inflammatory stimulation, nCD64 is rapidly upregulated and participates in immune regulation^2^, it can be used as a valuable marker for the diagnosis of infection, disease monitoring and prognosis evaluation of infectious diseases^3^. Human leukocyte antigen-antigen D related (HLA-DR) is a protein required by antigen-presenting cells to present antigens to T cells, and is expressed on monocytes, macrophages, and B lymphocytes. Increased HLA-DR expression reflects the activation of immune cells, while decreased HLA-DR expression indicates impaired antigen presenting capacity^4^. Monocyte HLA-DR (mHLA-DR) is critical for the initiation of adaptive immunity, and its low level is regarded as a marker of immunosuppression^5^. During infection, reduced mHLA-DR expression inhibits antigen presentation and immune response, leading to immune dysfunction. Its detection is helpful to assess disease severity and prognosis^6^, and monitoring mHLA-DR level also plays a good guiding role in immunoregulatory therapy.

The diagnostic and predictive value of elevated nCD64 and reduced mHLA-DR in infectious diseases is widely recognized. However, there are currently no standard to define the normal reference interval for these indicators, particularly regarding potential variations across different age groups. Without accurate reference value to evaluate infection status, clinicians may struggle to interpret results accurately, potentially leading to over-diagnosis or missed diagnosis. Therefore, it is necessary to establish age-specific reference intervals for nCD64 and mHLADR to support reliable clinical decision-making and improve patient management.

## 2. Material and Methods

### 2.1 Patients

Participant recruitment for this study was conducted from February 1, 2024, to February 1, 2025. A total of 320 healthy adults aged 14-100 years were initially recruited during physical examinations at Zigong First People’s Hospital. Exclusion criteria were abnormal blood routine indicators, liver and kidney function indicators, a history of genetic disease and obesity, recent illness, surgery, transfusion, and hospitalization. Finally, 285 cases were included in the study. The study protocol was approved by the Ethics Committee of the Zigong First People’s Hospital. Written informed consent was obtained from all participating adults. For participants who were minors, written informed consent was provided by their parents or legal guardians.The flow chart of this study is shown in Figure 1

**Figure 1:**
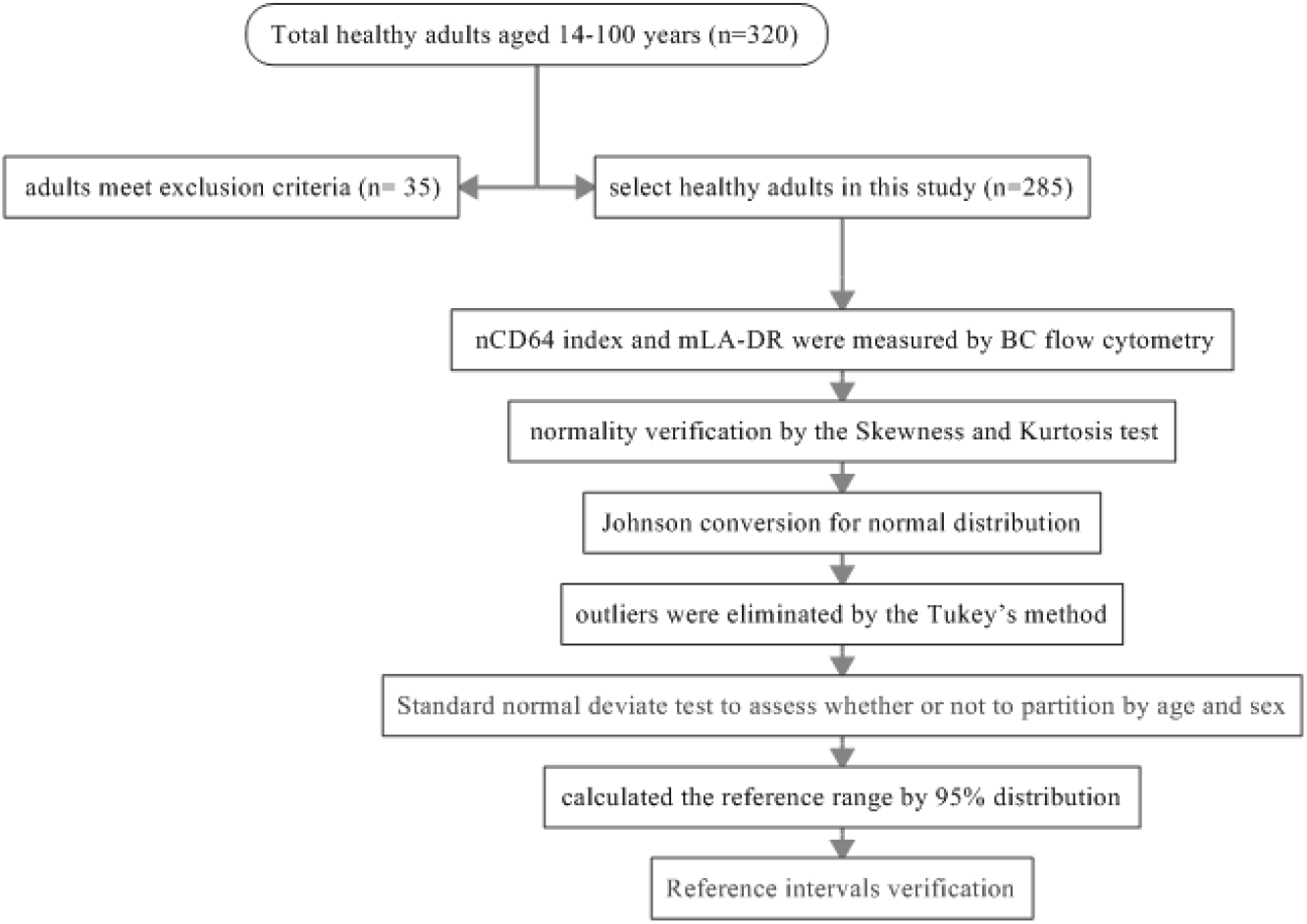
The flow chart of this study for establishing reference values.

### 2.2 Instruments and reagents

nCD64 index and mLA-DR were measured by BC flow cytometry on BC Navio system (American Beckman Coulter, California, USA). 50 μL of EDTA anticoagulated whole blood sample was collected and incubated with 5 μL anti-HLADR-FITC, 5 μL anti-CD64-PE, 5 μL anti-CD13-ECD, 5 μL anti-CD14-APC750 and 5 μL anti-CD45-PC7 at room temperature in the dark for 30 min. Hemolysin (2ml) for lysis, and then all samples were centrifuged for wash. The supernatant was removed, and 500μl of phosphate buffer was added. Data on 15,000 cells were acquired on a NAVIOS instrument (BC) and analyzed using Kaluza Analysis Software version 2.1 (BC, California, USA). The flow cytometer settings were prepared in accordance with the manufacturer’s instructions.

The scatter plot was drawn with side scatter (SSC) and pan-leucocyte marker CD45 to gate nucleated cells, Then, CD14/CD13-gating was used to isolate monocytes, granulocyte, and lymphocytes, all these cells was measured the median fluorescence intensity (MFI) of CD64, the corresponding indicators were Monocyte CD64-MFI (mCD64-MFI), Neutrophil CD64-MFI (nCD64-MFI), and Lymphocyte CD64-MFI (LyCD64-MFI), respectively. we calculate the nCD64 index according to the following formula: nCD64 index=[nCD64-MFI/Ly CD64-MFI]/[m CD64-MFI/ nCD64-MFI]^7^. The results of mHLA-DR were calculated by the percentage of HLA-DR cells in the monocytes (see Figure 2).

**Figure 2.**
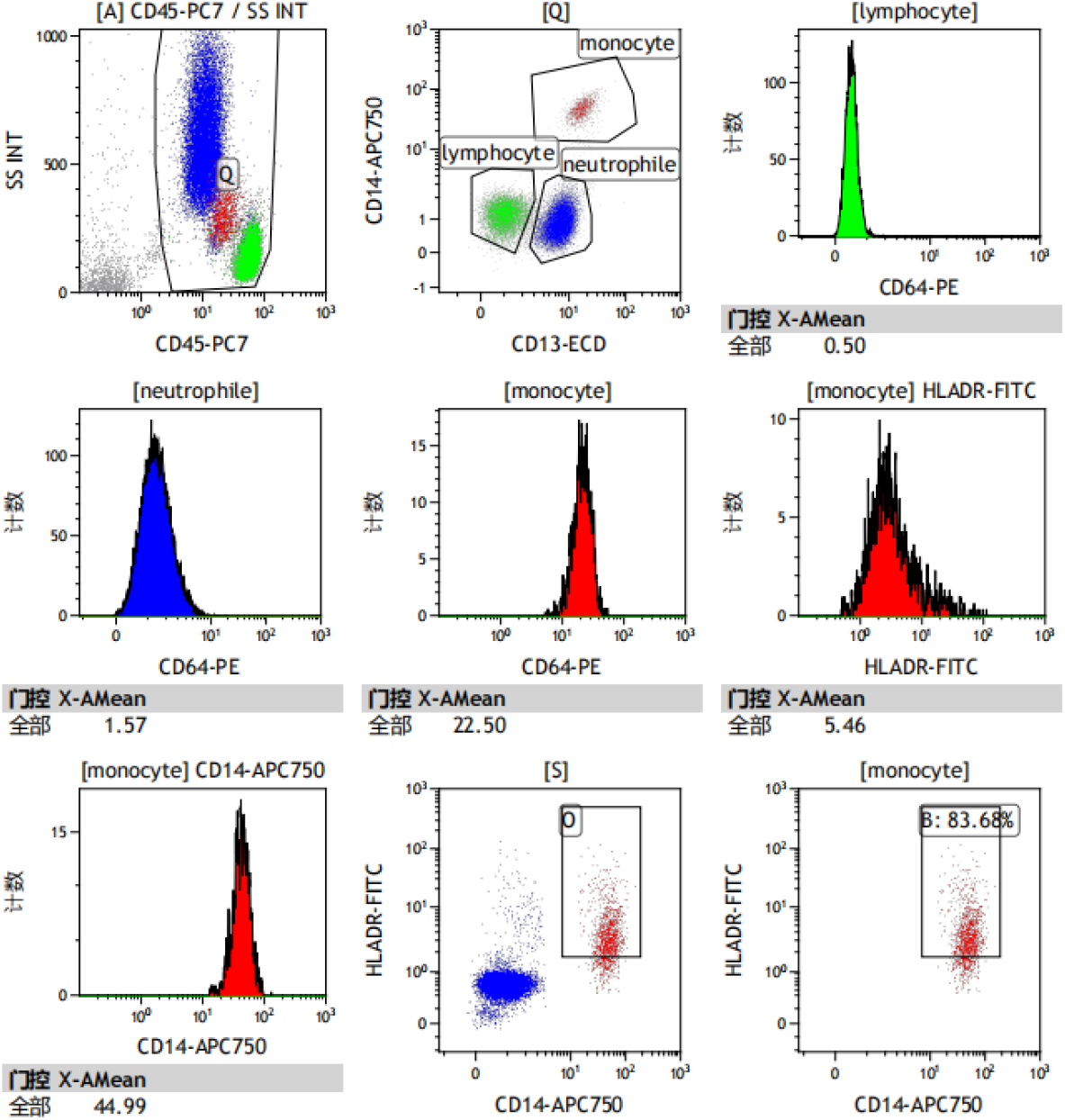
Schematic diagram of the flow cytometry analyses of nCD64 index and mHLA-DR.

### 2.3 Establishing reference values

First, data were analyzed for normality verification by the Skewness and Kurtosis test. If the standard deviation of the corresponding Skewness and Kurtosis were <1.96-times the Skewness and Kurtosis values, these data were judged as non-normal distribution that were transformed into approximately normal distribution by the Box-Cox. Second, the data outliers were eliminated by the Tukey’s method. If the data exceeded the upper and lower limits, they were regarded as outliers. Calculation formula of the upper limit was P75 plus 1.5-times the interquartile range (IQR), and that for lower limit was P25 minus 1.5-times the IQR. Further, all subjects were grouped into subgroups by sex and age. We performed the standard normal deviation test (Z-test) to evaluate the merging of reference intervals. Finally, we calculated the reference range by 95% distribution for the normal distribution and nonparametric data. The above methods were based on the methods in the CLSI EP28-A3C9 document published by the Clinical and Laboratory Standards Institute. Data were analyzed in accordance with the relevant guidelines.

### 2.4 Reference interval verification

Forty healthy adults in each age group were used for validating the obtained age-adjusted reference intervals. The ratio of individuals outside the reference intervals was calculated; if the ratio was <5%, the reference intervals were verified.

### 2.5 Statistical analysis

Statistical analyses were performed using Stata 15.0 Statistics (Stata Software Inc., La Jolla, CA, USA) and SPSS Statistics for Windows version 22.0 (SPSS Inc., Chicago, IL, USA). Continuous variables are reported as mean □ standard deviations (SD). The Kolmogorov–Smirnov test was used to assess normality. For non-normally distributed nCD64 index and mHLADR, comparisons between different age groups were made using nonparametric tests. The Kruskal– Wallis H test and Z-test were used to evaluate differences between age groups. A P < 0.05 was considered to be statistically significant.

## 3. Results

### 3.1 Distribution of data and elimination of outliers

The skewness and kurtosis of nCD64 index and mHLA-DR data obtained were 2.20 and 4.97, - 1.79 and 3.39, respectively, which is non-normally distributed by the Skewness-Kurtness test. After Box-Cox conversion, P value of nCD64 index was 0.737, and P value of mHLA-DR was <0.005 and 0.955. the skewness and kurtosis of nCD64 index and mHLA-DR data were −0.01 and −0.37, −0.01 and −0.16, respectively, showing an approximately normal distribution. The results before and after the transformation are shown in Table 1. We used the Tukey method to eliminate cases of outliers. See Figure 3 and Table 1 for detailed parameters before and after elimination.

**Table 1.** The data before and after outliers were eliminated by the Tukey method.

| Parameter | Before eliminate |  |  |  |  |  | After eliminate |  |  |  |  |  |
| --- | --- | --- | --- | --- | --- | --- | --- | --- | --- | --- | --- | --- |
|  | N | P<sub>25</sub> | P<sub>75</sub> | IQR | Max | Min | N | P<sub>25</sub> | P<sub>75</sub> | IQR | Max | Min |
| nCD64 | 285 | 0.18 | 0.54 | 0.36 | 2.33 | 0.06 | 269 | 0.17 | 0.44 | 0.27 | 1.05 | 0.06 |
| mHLA-DR | 285 | 89.04 | 97.43 | 8.39 | 99.85 | 35.86 | 265 | 90.43 | 97.55 | 7.12 | 99.85 | 78.53 |

**Figure 3.**
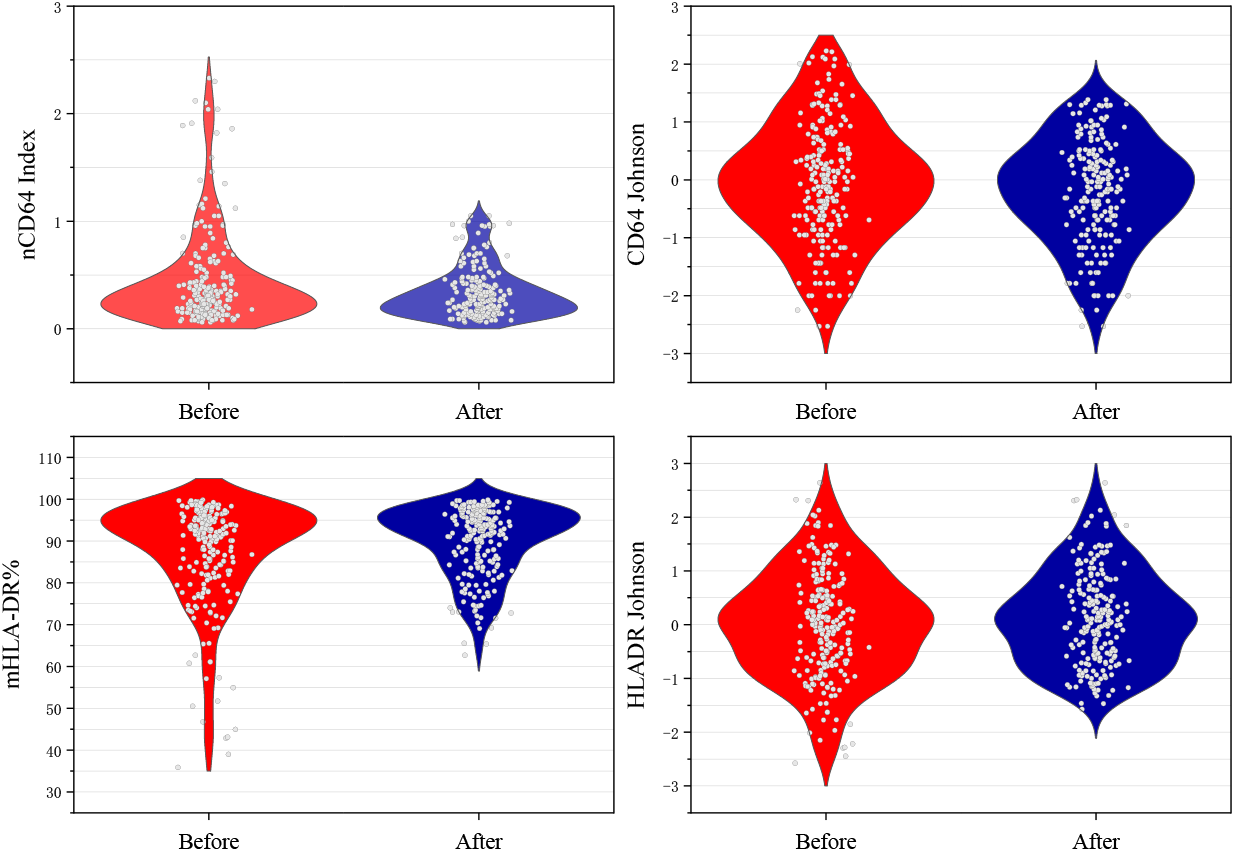
Violin plots for the data before and after outliers were eliminated by the Tukey method.

### 3.2 Establishment of the reference interval

We grouped the data by gender and age, all the subjects were grouped into subgroups according to gender (male and female) and age(14–50, and 51–100 years). The ANVOA analysis was used to evaluate whether age and gender were possible grouping factors. The standard normal deviate test (z-test) suggested by Harris and Boyd was used to assess whether or not to partition reference interval by the subclass. The results of ANVOA analysis indicated that gender and age might be not the dividing factors of mHLA-DR (p > 0.05), but the levels of nCD64 index were significantly different among age groups (p <0.05). further Z test results showed that it is necessary to set age-specific reference interval by subclass (Tables 2). The 95% distribution RI upper limit of nCD64 index and mHLA-DR after excluding outliers were calculated by the normal distribution method and the nonparametric method, respectively. For reference populations aged 14–100 years included in the study. The RI estimated by the normal distribution and the nonparametric method. All results were shown in Table 3.

**Table 2.** The nCD64 index results of pairwise comparison of age subclass by standard normal deviate test.

| Age groups (year) |  | F value | P value | Z | Z* |
| --- | --- | --- | --- | --- | --- |
| 14-50(n=124) | 51-100(n=145) | 26.881 | <0.001 | 5.225 | 4.775 |

$\bar{X}_1$ and $\bar{X}_2$ are the observed means of the two subgroups, $S_1$ and $S_2$ are the observed variances, and $N_1$ and $N_2$ are the number of reference values in each subclass, respectively. If the calculated Z exceeds $Z^*$ , they recommend partitioning.

**Table 3.**
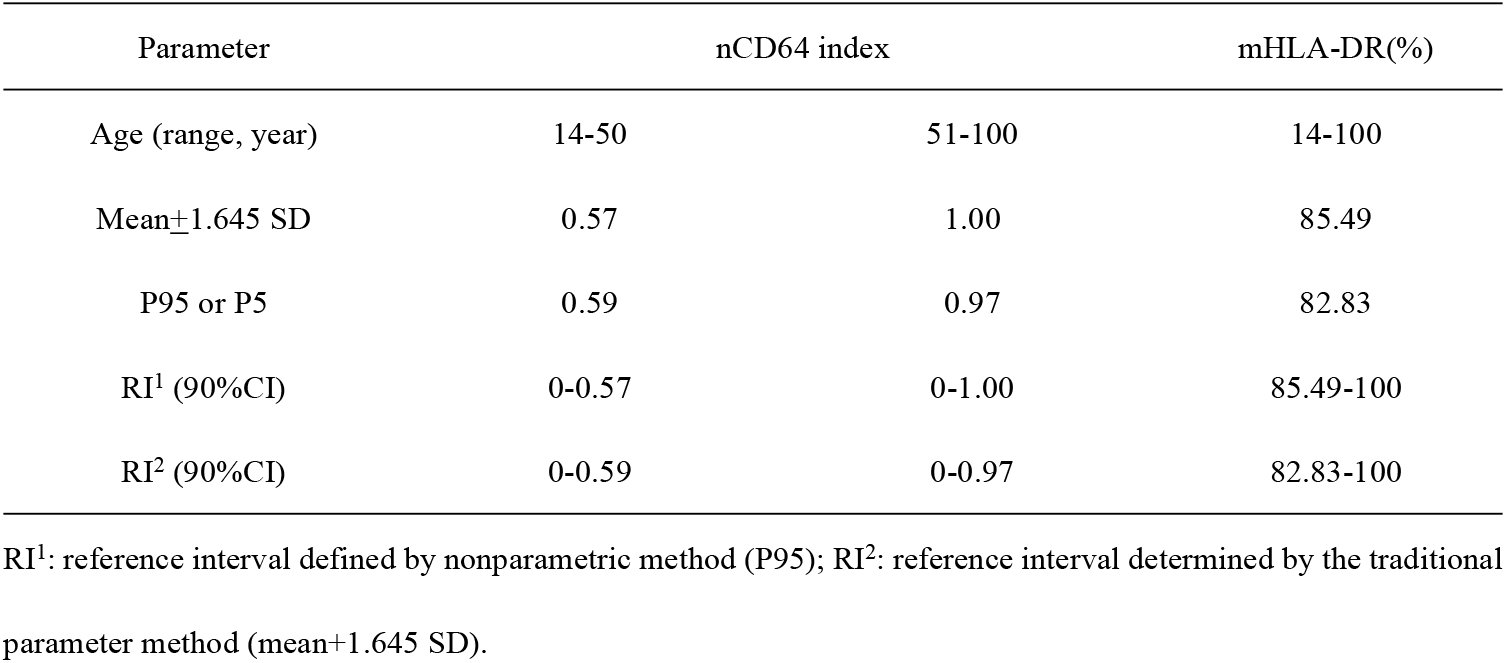
The nCD64 index and mHLA-DR reference interval calculate by parametric and nonparametric methods.

| Parameter | nCD64 index |  | mHLA-DR(%) |
| --- | --- | --- | --- |
| Age (range, year) | 14-50 | 51-100 | 14-100 |
| Mean±1.645 SD | 0.57 | 1.00 | 85.49 |
| P95 or P5 | 0.59 | 0.97 | 82.83 |
| RI<sup>1</sup> (90%CI) | 0-0.57 | 0-1.00 | 85.49-100 |
| RI<sup>2</sup> (90%CI) | 0-0.59 | 0-0.97 | 82.83-100 |

RI<sup>1</sup>: reference interval defined by nonparametric method (P95); RI<sup>2</sup>: reference interval determined by the traditional parameter method (mean+1.645 SD).

### 3.3 Verification of reference intervals

Some reference individuals were screened from the local reference population for verification (n = 40 per group). The factors before and during analysis were consistent with the reference interval in this study. The percentages of the nCD64 index and mHLA-DR subgroups from adults participating in verification of reference intervals are listed in Table 4. The results showed that all of results were above 95%, all passed verification.

**Table 4.**
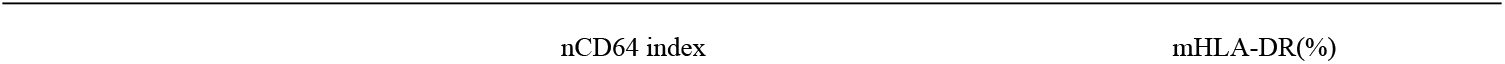
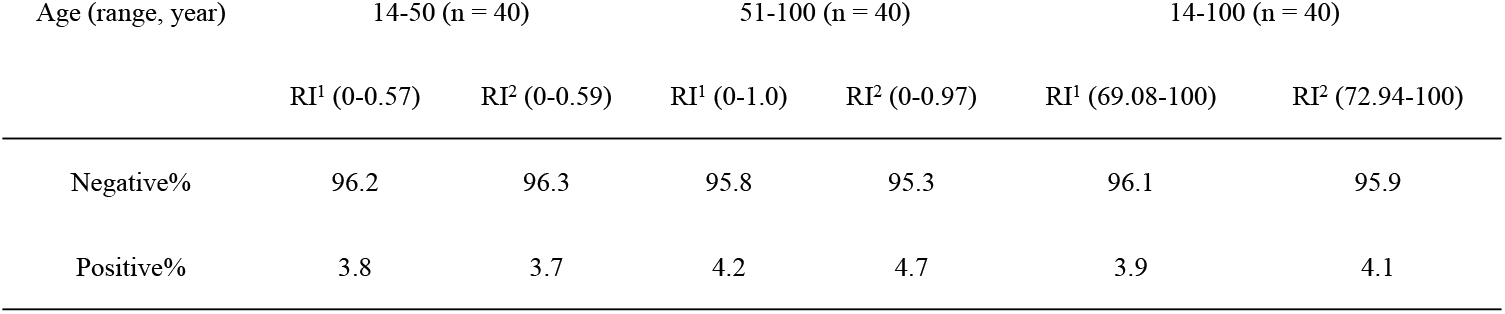
The comparison results of verify population by different nCD64 index and mHLA-DR reference intervals.

| Age (range, year) | 14-50 (n = 40) |  | 51-100 (n = 40) |  | 14-100 (n = 40) |  |
| --- | --- | --- | --- | --- | --- | --- |
|  | RI<sup>1</sup> (0-0.57) | RI<sup>2</sup> (0-0.59) | RI<sup>1</sup> (0-1.0) | RI<sup>2</sup> (0-0.97) | RI<sup>1</sup> (69.08-100) | RI<sup>2</sup> (72.94-100) |
| Negative% | 96.2 | 96.3 | 95.8 | 95.3 | 96.1 | 95.9 |
| Positive% | 3.8 | 3.7 | 4.2 | 4.7 | 3.9 | 4.1 |

### 3.4 Correlation analysis between Age and nCD64 index

In order to study whether there is a trend change of nCD64 index with the increase of age, we analyzed the correlation analysis between age and nCD64 index, CD64 MFI on lymphocyte, neutrophil and monocyte, we found that mCD64-MFI had a negative correlation with age (r = 0.3365, P < 0.00.01), and the others (nCD64-MFI and LyCD64-MFI) also have negative correlation with age(r = 0.2109, P = 0.0014; r = 0.3736, P < 0.0001, respectively). nCD64 index (CD64 MFI calculated value) have positive correlation with age (r = 0.3642, P < 0.0001), their interrelationships are shown in Figure 4. CD64 MFI interacted on lymphocytes, neutrophils and monocytes, and all of them increased with the increase of others. CD64 MFI on lymphocytes was positively correlated with CD64 MFI on neutrophils and on monocyte (r = 0.5585, P < 0.0001; r = 0.462, P < 0.0001, respectively), CD64 MFI on neutrophils was also positively correlated with CD64 MFI on monocyte (r = 0.4715, P < 0.0001), their interrelationships are shown in 3D graph (Fig 5).

**Figure 4.**
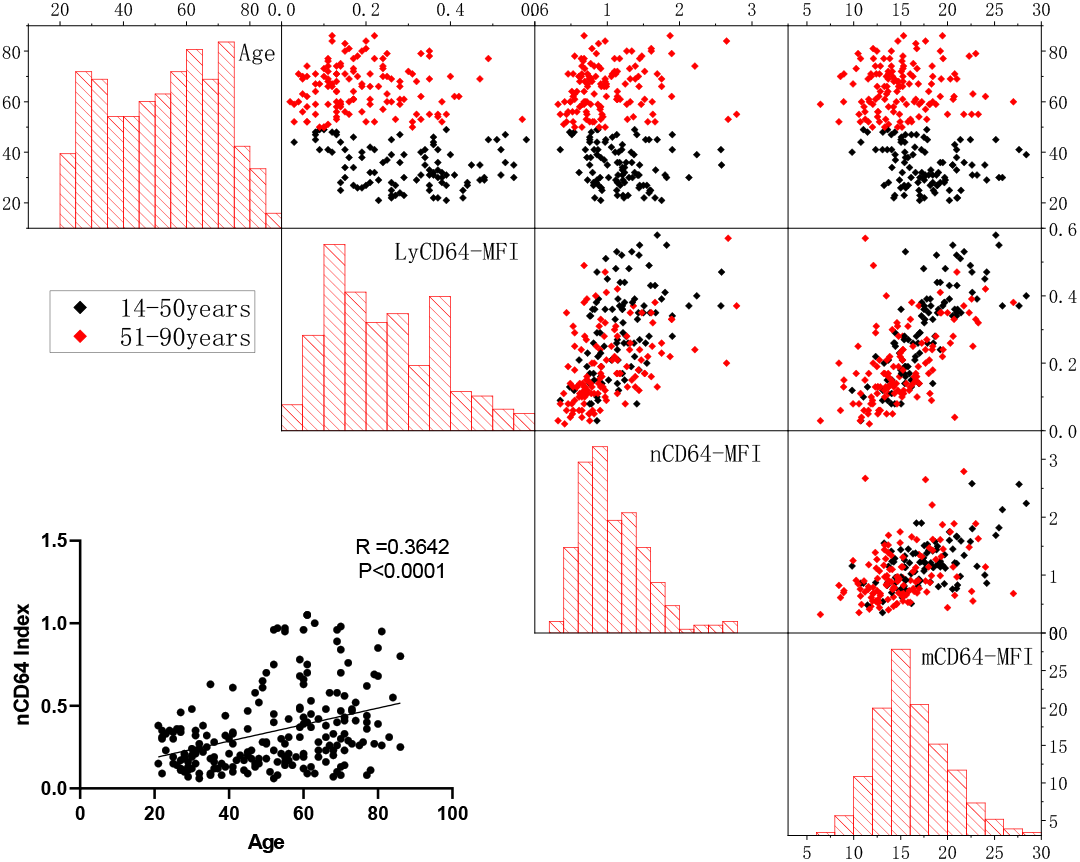
Scatter diagram for interrelationships between age, LyCD64-MFI, nCD64-MFI, mCD64-MFI and nCD64 index.

**Figure 5.**
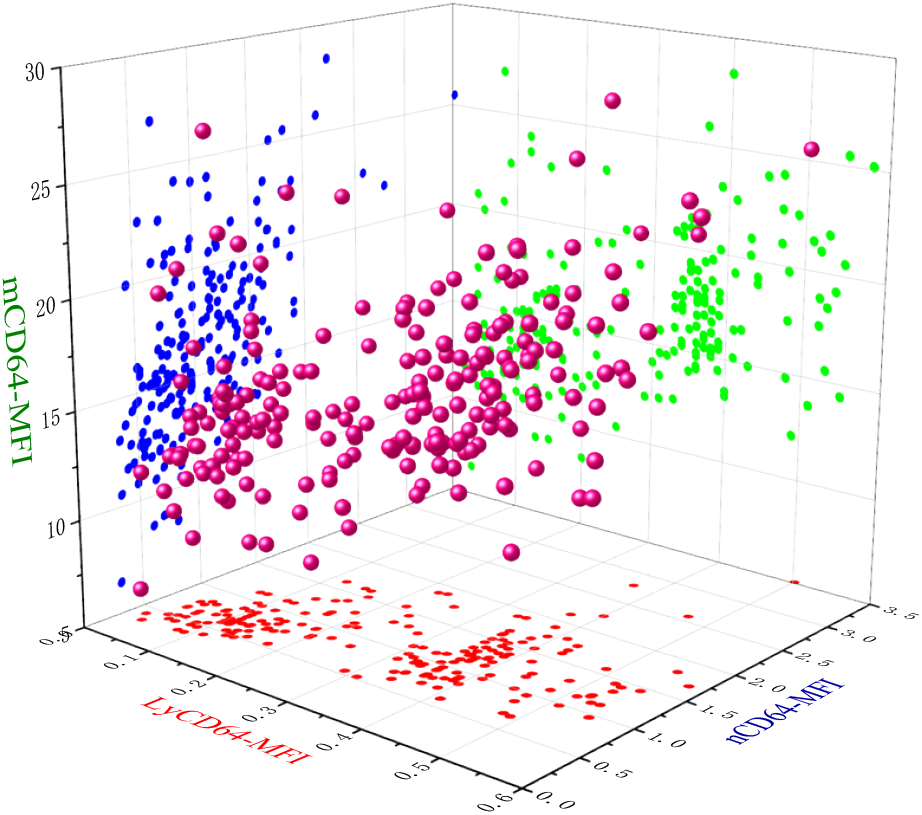
3D scatter diagram for interrelationships between LyCD64-MFI, nCD64-MFI and mCD64-MFI.

### 3.5 Correlation analysis between blood routine indexes and nCD64 index,mHLA-DR%

We further studied the correlation of blood routine indexes and derivative indexes (including PLR, SII, NLR, d-NLR, LMR) with nCD64 index and mHLA-DR%,we calculate the derivative indexes according to the following formula: PLR = number of platelet (PLT) / Absolute number of lymphocytes (LYM-abs), SII = number of PLT × Absolute number of neutrophils (NEU-abs) / LYM-abs, NLR = NEU-abs / LYM-abs, d-NLR = [White blood cells (WBC) - NEU-abs] / LYM-abs, LMR = LYM-abs / Absolute number of monocytes (Mono-abs). we found that no correlation between nCD64 index and mHLA-DR% and all the indicators of blood routine (P>0.05).

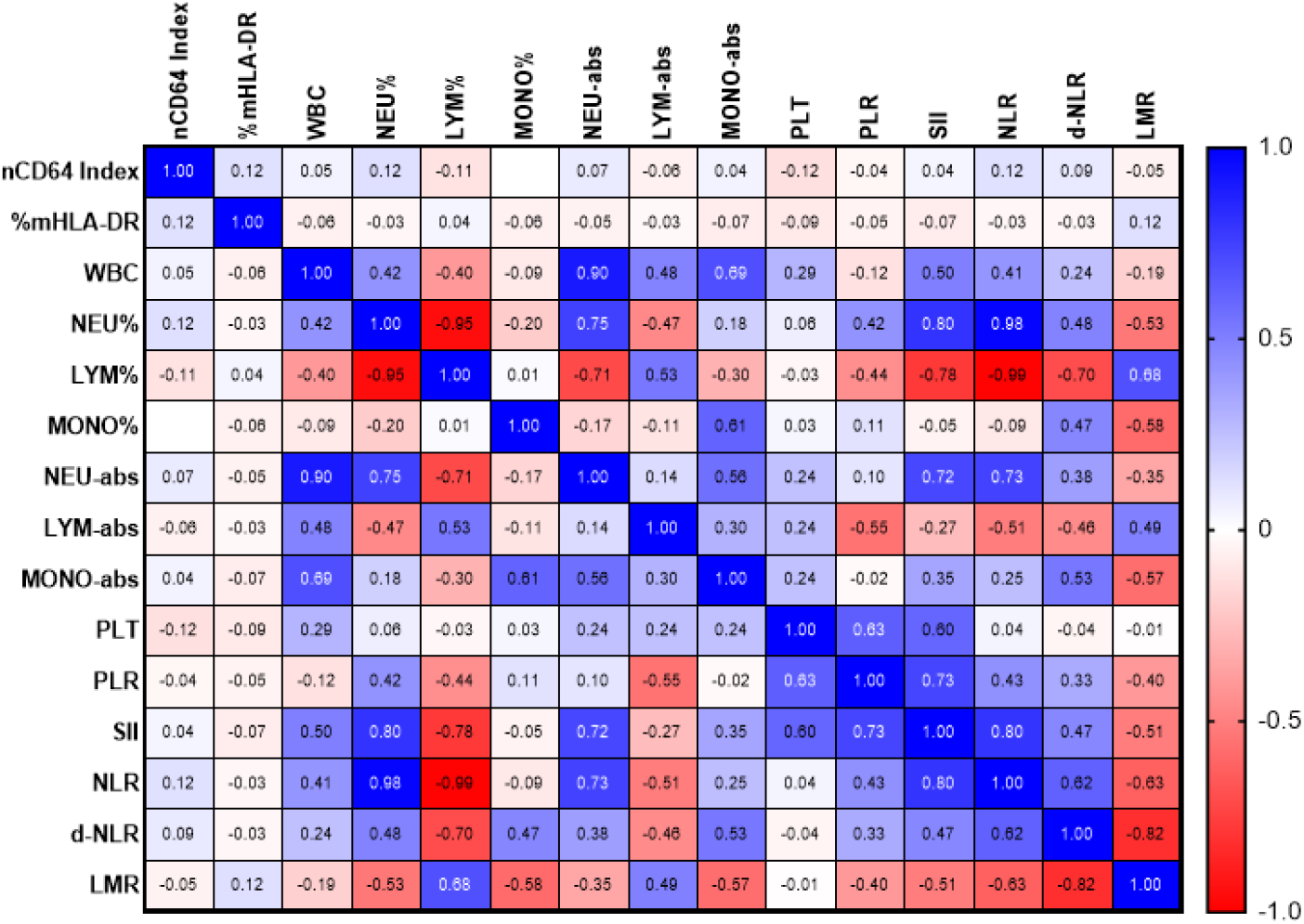

## 4. Discussion

CD64 is primarily expressed on the antigen-presenting cells. Under normal condition, neutrophil CD64 expression is low but increases rapidly within 4–6 hours after infection, serving as an early indicator of bacterial infection unaffected by hormones or antibiotics. CD64 detection can be performed using three methods: the percentage of CD64 positive cell, CD64 average fluorescence intensity and CD64 index. Among these, CD64 index minimizes the influence of external factors such as research objects and measuring instruments, offering higher sensitivity and specificity^8^. Therefore, this study used the nCD64 index to establish a reference interval for adults. The results showed that the mean value of nCD64 index was 0.57 in healthy individuals aged 14-50 years old, and 1.0 in those aged 51–100 years old. The nCD64 index gradually increased with age and showed a positive correlation with age (r = 0.3642, P < 0.0001). Many studies have indicated that the aging process is often accompanied by a state of chronic low-grade inflammation, which is described as inflammatory aging^9^. Unlike acute inflammation or infection, chronic low-grade inflammation is characterized by a long-term mild elevation of the representative inflammatory factors such as C-reactive protein and interleukin 6. Inflammatory aging may result from the accumulation of endogenous macromolecules or cellular debris, an increase in senescent cells and aging-related secretory phenotypes, declining immune function, alterations in microorganisms and their metabolites, and enhanced activity of the coagulation system^10^. Since nCD64 index is itself an indicator of inflammation, the findings of this study are consistent with the concept of inflammatory aging. However, there was no correlation between mHLA-DR and age in this study, indicating that although there was chronic low-grade inflammation with increasing age, it has minimal impact on the monocyte immune function. Specifically, there was neither immunosuppression nor enhanced immune activity in monocytes with age. Nevertheless, further studies with larger sample sizes are needed to strengthen the validity of these findings.

In this study, it was observed that the CD64 MFI on lymphocytes, neutrophils and monocytes exhibited a slight gradual increase with age. Further analysis showed that the CD64MFI values among these cell types interact, with the value in one cell type increasing in correlation with the fluorescence intensity of the other two. These findings indicate that CD64 MFI can be influenced by variables such as sample type, instrument voltage, and environmental conditions. Consequently, the observed trend of increasing CD64 MFI on lymphocytes and granulocytes with age is likely artifactual. Therefore, in order to minimize the impact of these external confounding factors, the CD64 index is currently recognized as the best method.

In order to verify whether the reference interval obtained in this study is applicable to other regions of China, we compared the range interval of CD64 index and mHLA-DR% for healthy control group in literature from other regions. As shown in Table 5, the upper value of CD64 index reference interval range from two different manufacturers of instruments were 0.59 and 1.21, respectively. In this study, the upper value of CD64 index reference interval was 0.57 for individuals aged 0–50 years and 1.0 for those aged 50–100 years, both of which fall within the range and are consistent with the reference interval for healthy people in other regions of china. According to the classification by reagent and instrument manufacturers, the upper reference value for CD64 index value obtained using Becton Dickinson instruments and reagents may be higher than that from Beckman Coulter systems. However, this observation requires further validation through direct comparison using the same samples analyzed with different instruments and reagents.

**Table 5.**
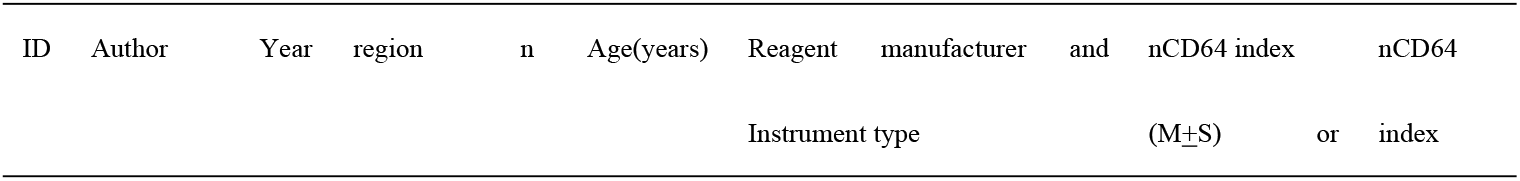
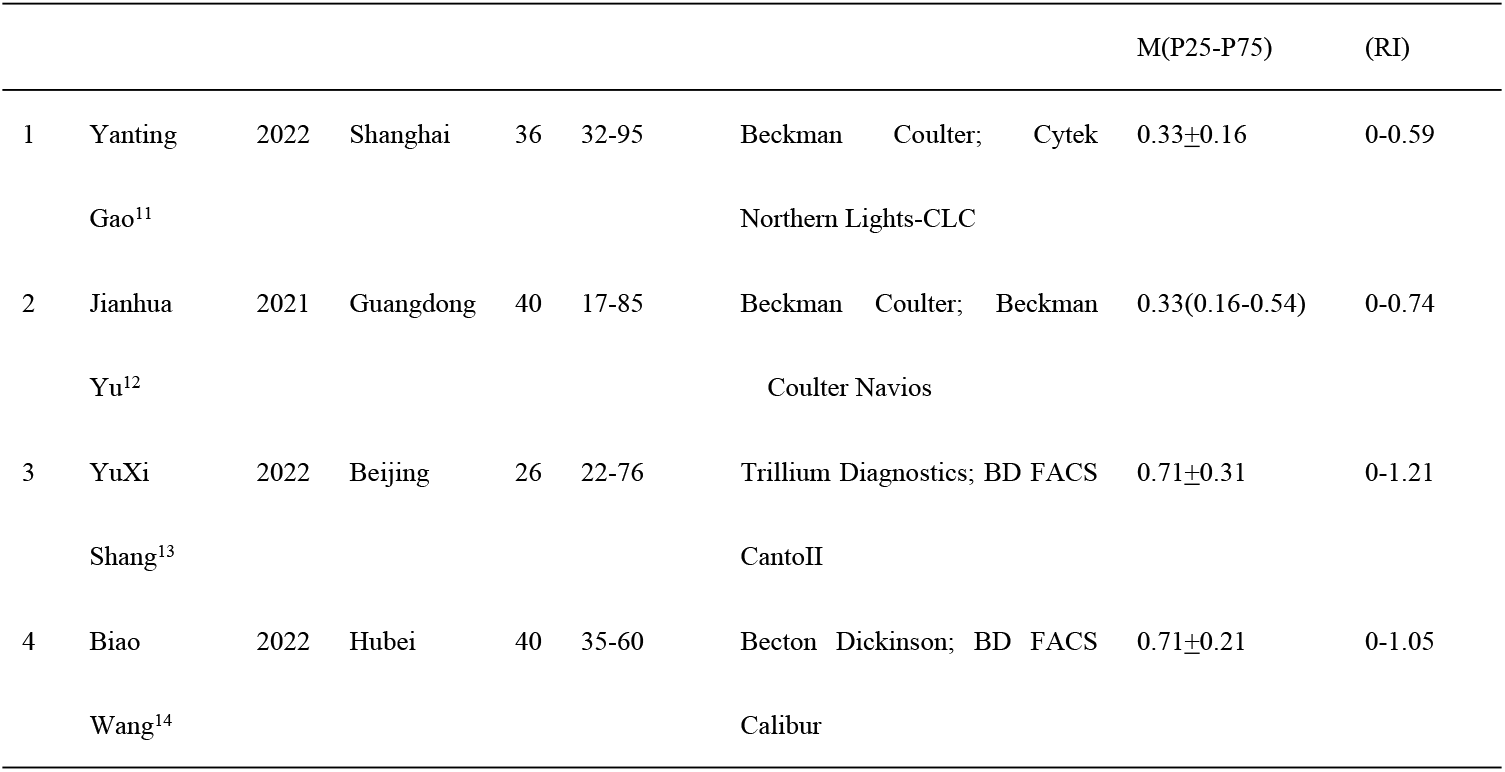
Reference intervals of nCD64 in adults: comparison among different regions in China.

Compared with the present study, the healthy reference range for mHLADR% reported by Jianhua Yu et al^12^ is lower (78.0-100), while that reported by Tianrong Zhou. et al^15^ is higher (90.4-100), which is relatively consistent with the reference range established in this study. However, both are notably higher than the values reported in a 2013 study by Shujun Zhou. et al^16^. HLADR is measured as the positive expression rate in monocyte and is susceptible to several factors, including fluorochrome itself, antibody binding capacity(ABC), and the fluorescence sensitivity of the instrument^17^. The characteristics of fluorochrome include its brightness and compatibility with the instrument’s excitation light. ABC refers to the number of specific high-affinity antibodies bound per cell under saturated staining conditions. The common interference factor is spectral overlap due to simultaneous staining with multiple antibodies on the same cells. The fluorescence sensitivity of the instrument includes both threshold and resolution^17^. The two flow cytometers may share the same threshold yet differ significantly in their ability to resolve dim populations. Therefore, it is recommended to establish distinct reference intervals for mHLA-DR% when using different instruments or fluorescent antibodies, but this needs further experimental verification to confirm.

The peripheral blood inflammatory indicators, such as systemic immune inflammatory index (SII), neutrophil to lymphocyte ratio (NLR), derived neutrophil to lymphocyte ratio (d-NLR), platelet to lymphocyte ratio (PLR), and lymphocyte to monocyte ratio (LMR), have been used as useful diagnostic or prognostic markers for various inflammatory diseases^18-20^. Accordingly, this study performed correlation analysis between these indicators and both the nCD64 index and mHLDR%. The results revealed no significant correlations between the inflammatory markers and either nCD64 index or mHLDR%, indicating that nCD64 index and mHLDR% may serve as independent risk indicators in inflammatory diseases.

## 5. Conclusion

The reference values of nCD64 index and mHLDR% established in this study are expected to support improved clinical evaluation and treatment of adult patients in China, A better understanding of age-related phenotypic changes is essential for the accurate identification of infectious diseases, particularly as these markers serve as critical independent risk indicators. Furthermore, future studies should prioritize extending this research to children under 14 years of age.

**Table 6.**
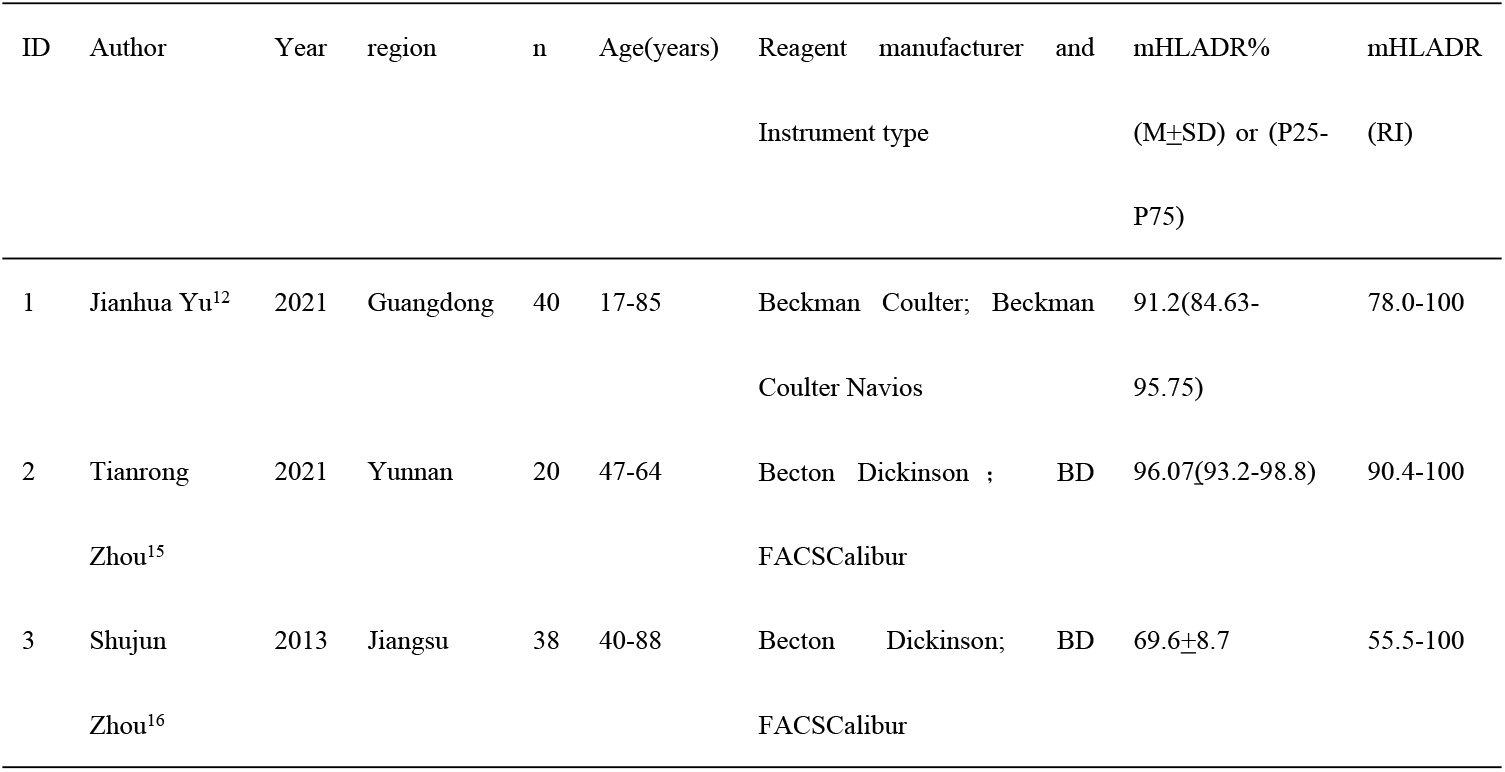
Reference intervals of mHLA-DR in adults: comparison among different regions in China.

## Data Availability Statement

The data supporting the findings of this study are available from the corresponding author upon reasonable request.

## Ethical approval

This study was approved by the Ethics Committee of the Zigong First People’s Hospital (NO.03202024).

## Author Contributions

Conceptualization, M.Y.; methodology, L.Z. and Y.L.; formal analysis, J.H. and L.Z; investigation, H.Z.; writing—original draft preparation, L.Z. and J.H.; writing—review and editing, Y.L., L.Z. and J.H.; visualization, M.Y.; supervision, M.Y. and Y.L.; project administration, M.Y. and Y.L. All authors have read and agreed to the published version of the manuscript.

## Acknowledgements

None

